# HDAC3 inhibition as a therapeutic strategy in T-cell acute lymphoblastic leukemia via the TYK2-STAT1-BCL2 signaling pathway

**DOI:** 10.1101/2025.02.27.25322993

**Authors:** Zhenyang Gu, Yuchen Liu, Yifan Jiao, Hao Wang, Lili Wang, Ning Le, Xiawei Zhang, Qingyang Liu, Yang Xu, Chunji Gao, Daihong Liu, Liping Dou

## Abstract

**Introduction:** Few advances have been made in treating T-cell acute lymphoblastic leukemia (T-ALL). Approaches targeting histone deacetylases (HDAC) have not been thoroughly investigated in T-ALL. However, the underlying molecular mechanism of HDAC inhibition remains to be fully elucidated.

**Objectives:** The study aimed to evaluate the clinical outcome of chidamide (an oral selective HDAC inhibitor for HDAC1, HDAC2, HDAC3, and HDAC10) in combination with chemotherapy in relapsed or refractory T-ALL and explore the underlying molecular mechanism of HDAC inhibition in T-ALL.

**Methods:** The clinical outcomes of 28 patients with relapsed or refractory T-ALL, who received chidamide in combination with chemotherapy were first evaluated. Transcriptomic analysis was used to identify pivotal signaling pathways of histone deacetylase inhibition in T-ALL cell lines. Short hairpin RNA-mediated inhibition, co-immunoprecipitation, and a series of functional assays were performed to verify the putative signaling pathways involved in cell lines, primary patient samples, and mouse models.

**Results:** Of the 28 patients, 16 achieved a complete response and three achieved a partial response. As an inhibitor of histone deacetylases, chidamide significantly suppressed the proliferation of T-ALL cells and induced apoptosis and cell cycle arrest in vitro. The protein level of HDAC3, but not of HDAC1, HDAC2, or HDAC10, was significantly inhibited by treatment with chidamide in T-ALL cell lines and primary human T-ALL cells. Moreover, the TYK2-STAT1-BCL2 signaling pathway was also substantially inhibited upon chidamide administration. Finally, overexpression of HDAC3 and TYK2 rescued the inhibitory effects of chidamide on T-ALL cells. HDAC3 bound to TYK2 and contributed to the activation of the TYK2-STAT1-BCL2 signaling pathway in T-ALL cells.

**Conclusion:** Our results highlight the effectiveness of the combination of chidamide and chemotherapy in the treatment of T-ALL patients and suggest that HDAC3 can act as a potential novel therapeutic target to inhibit the TYK2-STAT1-BCL2 signaling pathway in T-ALL.

**Novelty and Impact:** HDAC3 bound to TYK2 and contributed to the activation of the TYK2-STAT1-BCL2 signaling pathway in T-ALL cells.

HDAC3 can work as a potential alternative therapeutic target to inhibit the TYK2-STAT1-BCL2 signaling pathway in T-ALL.

## Background

T-cell acute lymphoblastic leukemia/lymphoma (T-ALL) is an aggressive malignancy caused by clonal proliferation of transformed thymic T-cell progenitor cells. It accounts for about 15% of pediatric ALL cases and 25% of adult ALL cases. Although there are distinct features in the demographics, clinical presentation, biology, and genetic landscape of T-ALL, it is still treated with chemotherapy similar to that used in B-cell acute lymphoblastic leukemia (B-ALL). Although cure rates have exceeded 75% in pediatric patients and about 50% in adult patients with intensified chemotherapy, the prognosis of patients with primary resistant or relapsed disease remains poor[1–3]. Furthermore, high toxicity or even mortality was also prominent in patients using intensified pediatric-based regimens[4–6]. Unlike the great success in the treatment of B-ALL, which incorporates inotuzumab, blinatumomab, and CAR-T therapy, few advances have been made in the treatment of T-ALL[7, 8]. Thus, novel effective targeted therapeutic strategies are urgently needed for patients with T-ALL[9, 10].

The genetic defects of T-ALL have been thoroughly investigated[11]. Many drugs have been developed against these oncogenic drivers belonging to different signaling pathways, such as those in NOTCH signaling, the cell cycle, and hematopoiesis[12]. However, to date, clinical outcomes of these drugs are unsatisfactory, somewhat reflecting the complex nature of this disease. More importantly, as in any other type of cancer, great progress on the epigenetic alterations in T-ALL has also been made[13]. Alterations of histone deacetylases (HDACs) have also been found in T-ALL[14]. Therefore, deregulated alterations in HDAC may provide an interesting therapeutic target for T-ALL. HDAC inhibition can induce apoptosis and growth arrest in many hematological malignancies and has been tested both in vitro and in vivo[15–18]. Based on this rationale, several classes of HDAC inhibitors have been developed and found to have potent antitumor effects with remarkable tumor specificity, both in preclinical and clinical studies. Chidamide, which can specifically inhibit HDAC1, HDAC2, HDAC3, and HDAC10, has also been evaluated as a monotherapy or combination therapy in clinical trials[19–23]. Mechanistic studies indicated that the molecular processes underlying the inhibition of HDAC were much broader and more complicated than originally understood[24]. Previous studies have demonstrated that several signaling pathways, such as the WNT/β-catenin and NOTCH/MYC signaling pathways, mediate the anti-tumor effects of HDAC inhibition in T-ALL[25–27]. Given the pleiotropic effects of HDAC inhibition, it is more reasonable to believe that it may produce more powerful effects if it was used in combination with chemotherapy. However, the underlying molecular mechanism of HDAC inhibition remains to be fully elucidated.

Here, our study found that the HDAC3-TYK2-STAT1-BCL2 signaling pathway was activated in T-ALL cells. Chidamide, when combined with chemotherapy, improved the survival of patients with relapsed/refractory (R/R) T-ALL. We further found that HDAC3 bound to TYK2 and contributed to the activation of the TYK2-STAT1-BCL2 signaling pathway in T-ALL cells. Our study indicated that HDAC3 is a potential novel therapeutic target that inhibits the TYK2-STAT1-BCL2 signaling pathway in T-ALL.

## Materials and methods

### Clinical study

Patients who were diagnosed with R/R T-ALL and T-cell lymphoblastic lymphoma (T-LBL) and who received chidamide during salvage chemotherapy were recruited between May 2016 and June 2019 at the First Medical Center of the Chinese PLA General Hospital. This retrospective study was designed to evaluate the clinical efficacy and safety of combining chidamide with chemotherapy for patients with R/R T-ALL. The following inclusion criteria were used: ① patients whose diagnosis met the criteria of T-ALL or T-LBL according to the 2016 edition of the WHO classification of precursor lymphoid neoplasms; ② patients with primary refractory disease after induction therapy; ③ patients who relapsed within 12 months after first complete remission; ④ patients who were refractory to or relapsed after at least one salvage treatment; and ⑤ patients with an Eastern Cooperative Oncology Group (ECOG) score ≤ 2. Patients were excluded in this study if they had severe organ dysfunction (heart, kidney, liver, or other organs). The criteria of ETP was defined as previously[28]. Our study was reviewed and approved by the Ethics Committee of the Chinese PLA General Hospital. All patients signed consent forms for the collection and use of their cells and clinical demographic data.

Chidamide (30 mg) was orally administered twice a week for a total of four doses during the first 2 weeks of the combined salvage chemotherapy. The chemotherapy regimen was selected according to the patient’s pretreatment history and physical status (see Table S1). The choice of salvage chemotherapy was determined according to the patient’s physical status, previous treatment history and response, and the discretion of the oncologist in charge. Response assessments, including the evaluation of bone marrow, peripheral blood, and extramedullary lesions, were usually performed on the 28th day of each course of treatment or until hematopoietic recovery. Complete remission (CR) was defined as the percentage of bone marrow blasts dropped to be less than 5% and the resolution of extramedullary masses. A partial response (PR) was defined as the percentage of bone marrow blasts dropped to 6–25% and (or) more than 50% reduction of the tumor mass. Progressive disease (PD) was defined as the number of blast cells in the bone marrow or peripheral blood increased by more than 25%, more than 25% increase of the tumor mass, or the onset of a new tumor mass. No response (NR) was defined as the failure to meet the criteria of a PR or PD. The overall response rate (ORR) was defined as the CR plus PR. Adverse effects were evaluated according to the National Cancer Institute Common Toxicity Criteria.

### Reagents and cell culture

Chidamide was obtained at no cost from Chipscreen Biosciences Ltd. (Shenzhen, China). RPMI 1640 medium, fetal bovine serum, and dimethyl sulfoxide (DMSO) were all purchased from Gibco Company (USA). All antibodies for Western blotting were purchased from Cell Signaling Technology (Danvers, MA, USA), except for TYK2 and p-STAT1, which were from Abcam (Cambridge, UK). T-ALL cell lines, including Jurkat and MOLT-4, were obtained from ATCC (Manassas, VA, USA). Primary T-ALL patient cells were isolated from bone marrow. Bone marrow samples were first diluted with an equal volume of PBS solution. The diluted mixture was spread over lymphocyte separation fluid (Lymphoprep 07801, Stemcell, USA) in a centrifuge tube and centrifuged at 2800 rpm for 25 minutes at 25°C. Then, the mononuclear cells between the plasma layer and separation fluid were collected. Jurkat, MOLT-4, and primary cells were maintained in RPMI 1640 supplemented with 10% fetal bovine serum, 100 μg/mL penicillin, and 10 μg/mL streptomycin at 37°C in a humidified atmosphere containing 5% CO_2_.

### Cell proliferation assays

Jurkat and MOLT-4 cells (3×10^4^ cells) were seeded in 96-well plates (each well contained 100 μL of cell suspension). Cells were incubated with different concentrations of chidamide for 24, 48, or 72 h. Then, 10 μL of CCK-8 reagent (Dojindo Laboratories, Kumamoto, Japan) was added to each well and incubated for an additional 2–3 h. The OD values of each well were measured by a microplate reader at 450 nm.

### Cell cycle analysis

The chidamide-treated cells were washed twice with cold PBS and fixed with cold 70% ethanol overnight. After fixation, the cells were washed with cold PBS and stained in PBS supplemented with 100 µg/mL RNase A and 100 µg/mL PI (Sigma) for 15 min. Samples were processed on a flow cytometer with a 488-nm laser and then analyzed with a MoFlo MLS sorter (Dako, Fort Collins, CO).

### Cell apoptosis assay

Cells were collected after being treated with chidamide for the indicated time. They were washed with ice-cold PBS and incubated in 200 μL of binding buffer containing 10 μL of PI and 5 μL annexin-V-FITC (Sigma) for 15 min at room temperature in the dark. Then, the stained cells were analyzed on a flow cytometer. All data were analyzed by FlowJo version 10.

### HDAC activity analysis

HDAC activity was measured by a Colorimetric HDAC Activity Assay kit (BioVision, San Francisco, USA). Each reaction was performed in a 100-μL reaction system that contained 50 μg of protein extracted from cells, 10 μL of HDAC assay buffer, and 5 μL of an HDAC colorimetric substrate. The reaction mixture was incubated at 37°C for 1 h and then was stopped by adding 10 μL of lysine developer. The reaction system was then incubated at 37°C for another 30 min. HDAC activity was measured on a microplate reader (SpectramMax M5) at 405 nm.

### RNA sequencing and analysis

Chidamide-treated and DMSO-treated Jurkat and MOLT-4 cells were collected and washed twice with ice-cold PBS. Total RNA was extracted using TRIzol reagent (Invitrogen, USA) following the manufacturer’s instructions. RNA sequencing was performed on a Genome Analyzer IIx (Illumina, San Diego, CA, USA). Expression levels were calculated with the RPKM method. The distribution of gene expression was analyzed.

### Real-time PCR

Total cellular RNA was extracted with TRIzol according to the manufacturer’s instructions. cDNA was synthesized using a PrimeScript™ RT reagent kit (Takara). Real-time PCR (RT-PCR) assays were performed using a KAPA SYBR FAST q-PCR Master Mix (2x) kit. Primers for target genes are listed in Table S2. The relative quantification of target genes was performed using 2-ΔΔCt. Each sample was analyzed in triplicate.

### Plasmids and transfection

*HDAC3* and *TYK2* plasmids and their shRNA constructs were cloned into the lentiviral vector pLKO.1-puro. *HDAC3* and *TYK2* plasmids and their corresponding control plasmid were purchased from Genechem. The *HDAC3* shRNA plasmid (sc-35,538-SH) and *TYK2* shRNA plasmid (sc-27,545-SH) were from Santa Cruz. Jurkat and MOLT4 cells were transfected with lentivirus-containing medium according to the manufacturer’s protocol. Then, cells were selected using antibiotics. They were then expanded and subjected to RT-PCR and Western blot.

### Western blot and co-immunoprecipitation analysis

After cells were collected and washed twice with ice-cold PBS, they were lysed in RIPA buffer that contained phosphatase inhibitors and protease inhibitors. Equal amounts of proteins were separated by SDS-polyacrylamide gel electrophoresis (SDS-PAGE) and transferred onto polyvinylidene difluoride (PVDF) membranes (Millipore). The PVDF membranes were blocked with 5% non-fat dry milk for 1 h at room temperature and then were incubated overnight at 4°C with primary antibodies. Each membrane was incubated with a secondary antibody according to the manufacturer’s instructions. Then, target proteins were detected with enhanced chemiluminescence kits (Millipore, Billerica, MA, USA). For co-immunoprecipitation, about 0.5–1 mg of protein lysate of Jurkat cells was immunoprecipitated by TYK2 and HDAC3 antibodies using co-immunoprecipitation reagents (ThermoFisher, #88804). The immune complex was subjected to immunoblotting with antibodies against HDAC3, TYK2, and p-TYK2.

### In vivo study

Xenograft experiments were established in NOD/SCID immunodeficient mice as previously described[29]. Athymic mice were maintained in laminar flow air conditioning cabinets and fed with a standard diet of laboratory rodent food and water under specific pathogen-free conditions. Jurkat cells (1×10^7^ cells in 200 μL of PBS) were injected subcutaneously into the lateral flanks of mice. Tumor volumes and weights were measured every other day. When the tumors approached 50 mm^3^ in size, the tumor-bearing mice were randomly assigned to different groups and treated with chidamide, deucravacitinib (a highly selective inhibitor of tyrosine kinase 2, MedChemExpress, China, BMS-986165), a combination of the two drugs, or normal saline as a control. The mice in the chidamide-treated group were intragastrically administered 5 mg/kg chidamide in PBS three times a week for 2 weeks. The mice in the deucravacitinib-treated group were intragastrically administered 30 mg/kg chidamide in PBS twice a day for 2 weeks. The mice in the control group were treated with both PBS and normal saline as a control. The mice in the combined therapy-treated group received intragastric instillation of both chidamide and deucravacitinib. The mice were sacrificed on the 21st day after inoculation. Tumors were measured and collected for further analysis. All protocols were approved by the Experimental Animal Ethics Committee of the Chinese PLA General Hospital. All animal experiments were performed in accordance with the international and institutional rules of the Institutional Animal Care and Use Committee (IACUC).

### Statistical analysis

Data were expressed as mean ± standard deviation (SD). Patient survival analysis was calculated using the Kaplan-Meier method with the log-rank (Mantel Cox) test. Statistical analysis was performed using SPSS 25.0 software and GraphPad Prism 9.0. The Mann– Whitney test or Student’s *t* test was used to perform comparisons when necessary. Differences were considered statistically significant when a two-tailed P-value was < 0.05.

## Results

### Combination of chidamide with chemotherapy treatment for patients with relapsed/refractory T-ALL

To evaluate the clinical efficacy of the combination therapy of chidamide and chemotherapy, 28 patients with R/R T-ALL received salvage chemotherapy and chidamide between May 2016 and June 2019 at the Chinese PLA General Hospital. The clinical characteristics of these patients are shown in Table 1. Nineteen patients were male and nine were female. The median age of these patients was 26 (range, 13–63) years. Six patients were diagnosed with T-LBL and the remaining 16 patients were diagnosed with T-ALL. Ten patients met the criteria of the ETP immunophenotype. Before salvage treatment, 12 (43%) patients had relapsed and 16 (57%) patients had refractory disease. The previous treatments of these 28 patients are listed in Table S2.

**Table 1.**
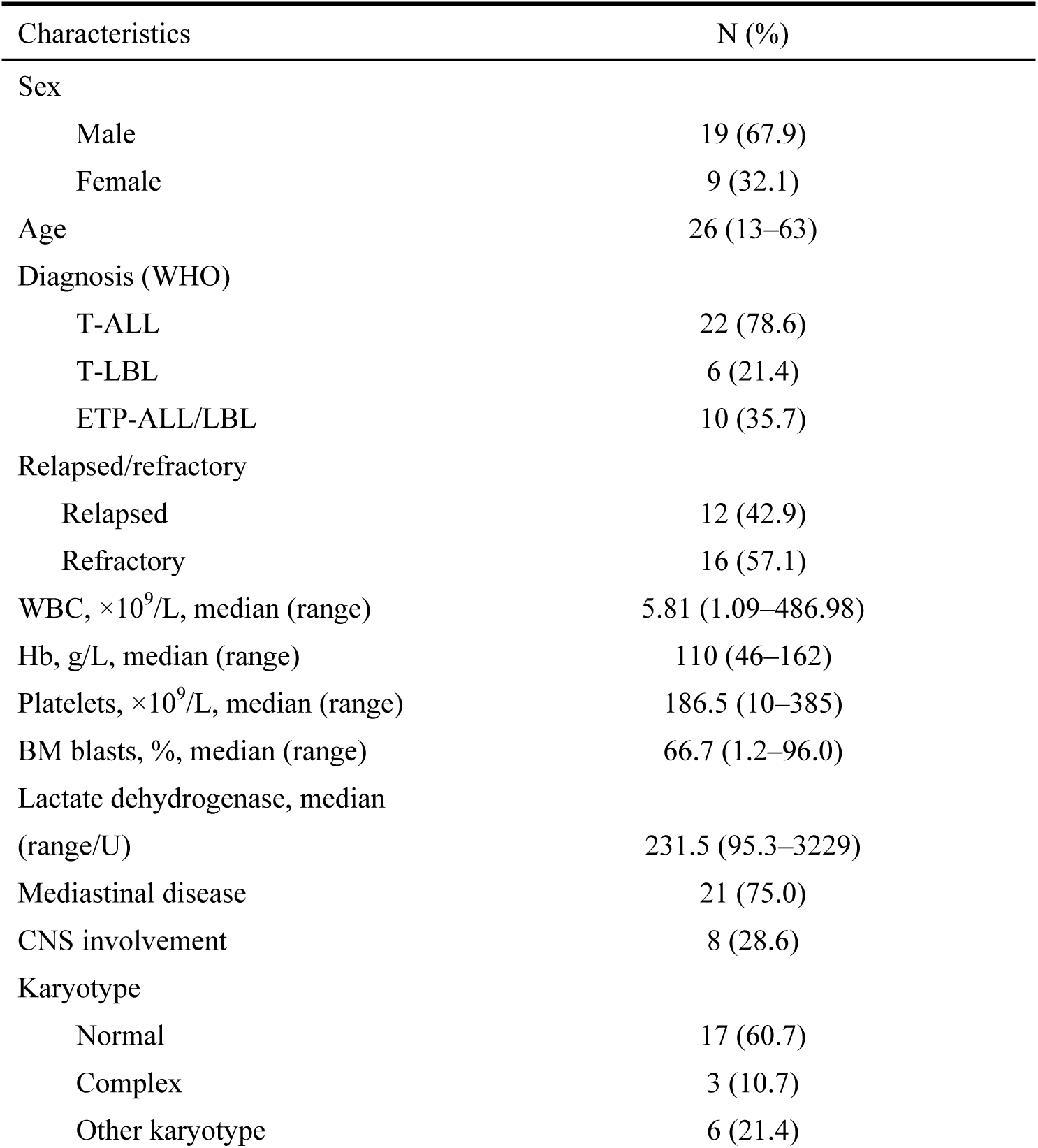
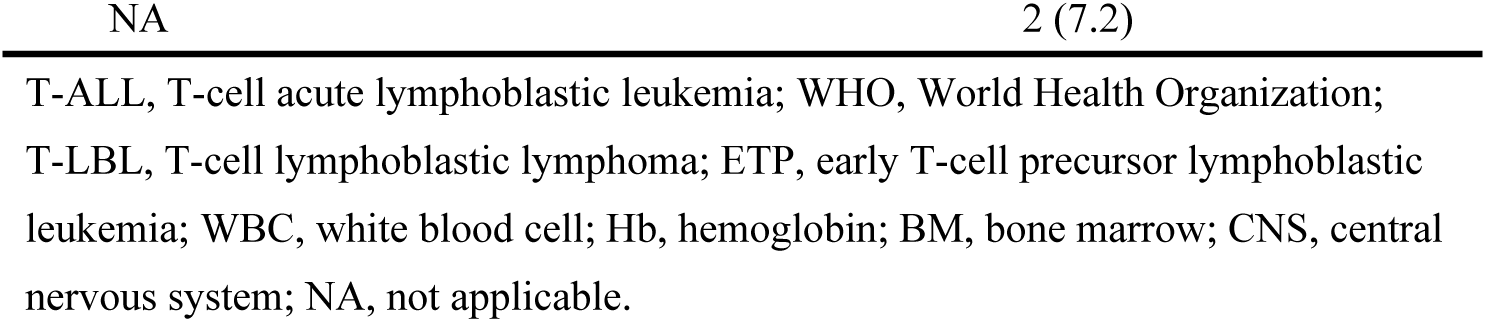
Characteristics of 28 patients with refractory/relapsed T-ALL.

During the study period, 28 patients received at least one course of salvage therapy. The salvage reinduction regimen of each patient and their response are shown in Table S3. Sixteen patients achieved CR and three patients achieved PR. The overall response and CR rate were 68% and 57%, respectively. The CR rate for patients with ETP-ALL was 70% (7/10). Twenty-one patients underwent allogeneic hematopoietic stem cell transplantation (allo-HSCT). Sixteen patients were in CR before allo-HSCT. Five patients with progressive disease received allo-HSCT as salvage therapy. Median follow-up for survivors was 56.5 (32–70) months. The OS rates at 1 and 3 years were 56.3% and 37.5%, respectively (Fig. 1A).

**Fig. 1.**
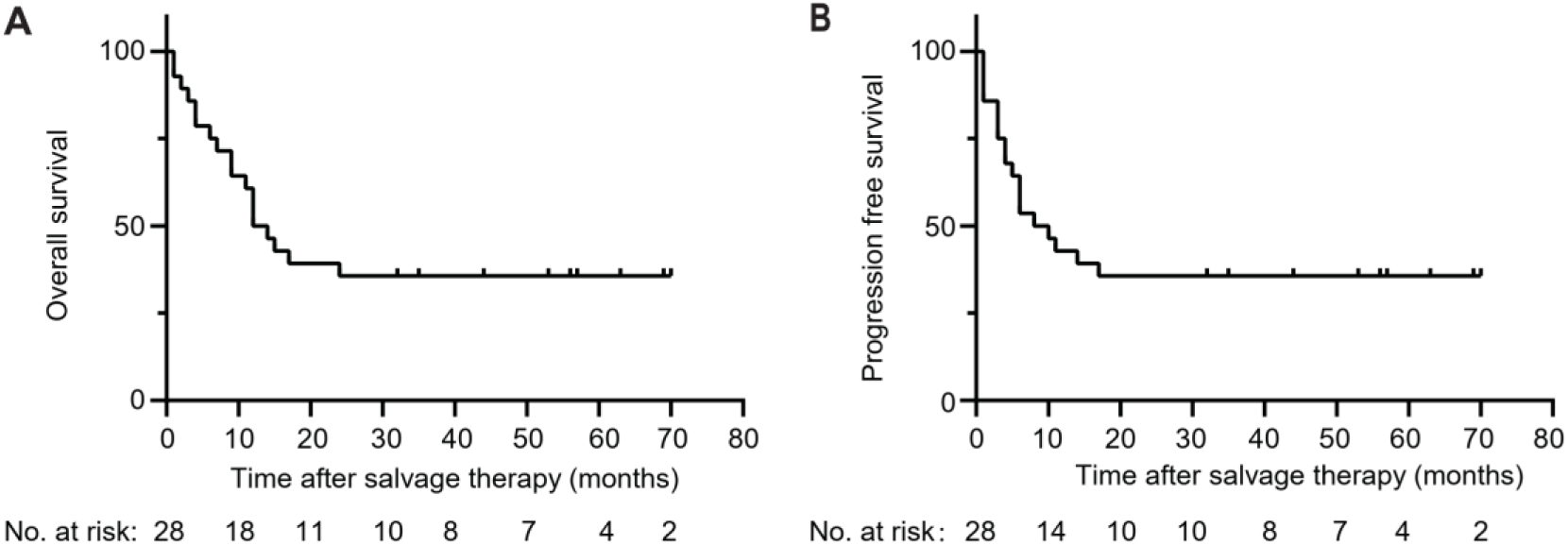
Kaplan-Meier estimates of survival rates of patients with refractory/relapsed T-ALL following combination therapy of chidamide and a chemotherapy regimen. **A** OS. **B** Progression-free survival. T-ALL, T-cell acute lymphoblastic leukemia; OS, overall survival.

The progression-free survival rates at 1 and 3 years were 42% and 27%, respectively (Fig. 1B). The occurrence of adverse events is listed in Table S4. All patients developed Grade III/IV bone marrow suppression. Febrile neutropenia was the most frequent event, which occurred in 14 cases (50%). Severe septic shock occurred in two cases. Other adverse events (more than one case) included grade I drug-induced liver injury (five cases), oral mucositis (five cases), diarrhea (three cases), hypofibrinogen (three cases), and pneumonitis (three cases).

### Chidamide significantly inhibited the proliferation of T-ALL cells and induced apoptosis and cell cycle arrest in vitro

The CR rates of salvage chemotherapy in R/R T-ALL range from 30% to 50%[30]. Therefore, improving the CR rate is a very critical step for this group of patients. The findings in this study, in which the CR rate was as high as 57% with the combined regimen, prompted us to investigate the underlying mechanism of chidamide-based therapy in T-ALL. Jurkat and MOLT-4 cells were treated with 0.3, 1, or 3 μM chidamide for 24, 48, or 72 h. As shown in Fig. 2A, the proliferation of both cell lines was significantly inhibited by chidamide in a dose- and time-dependent manner. Then, we determined whether chidamide induced apoptosis in T-ALL cells. Compared with control cells, the percentage of apoptotic cells increased when the concentration of chidamide increased (Fig. 2B, and Figure S1A, B). Furthermore, we tested whether chidamide induced cell cycle arrest in T-ALL cells. Consistent with a previous study[31], chidamide treatment increased the percentage of cells in the G0/G1 phase while decreasing the percentage of cells in the S phase (Fig. 2C and Figure S1C). The effect of chidamide on apoptosis and the cell cycle was also concentration- and time-dependent. Taken together, these data indicated that chidamide significantly inhibited the proliferation of T-ALL cells in vitro.

**Fig. 2.**
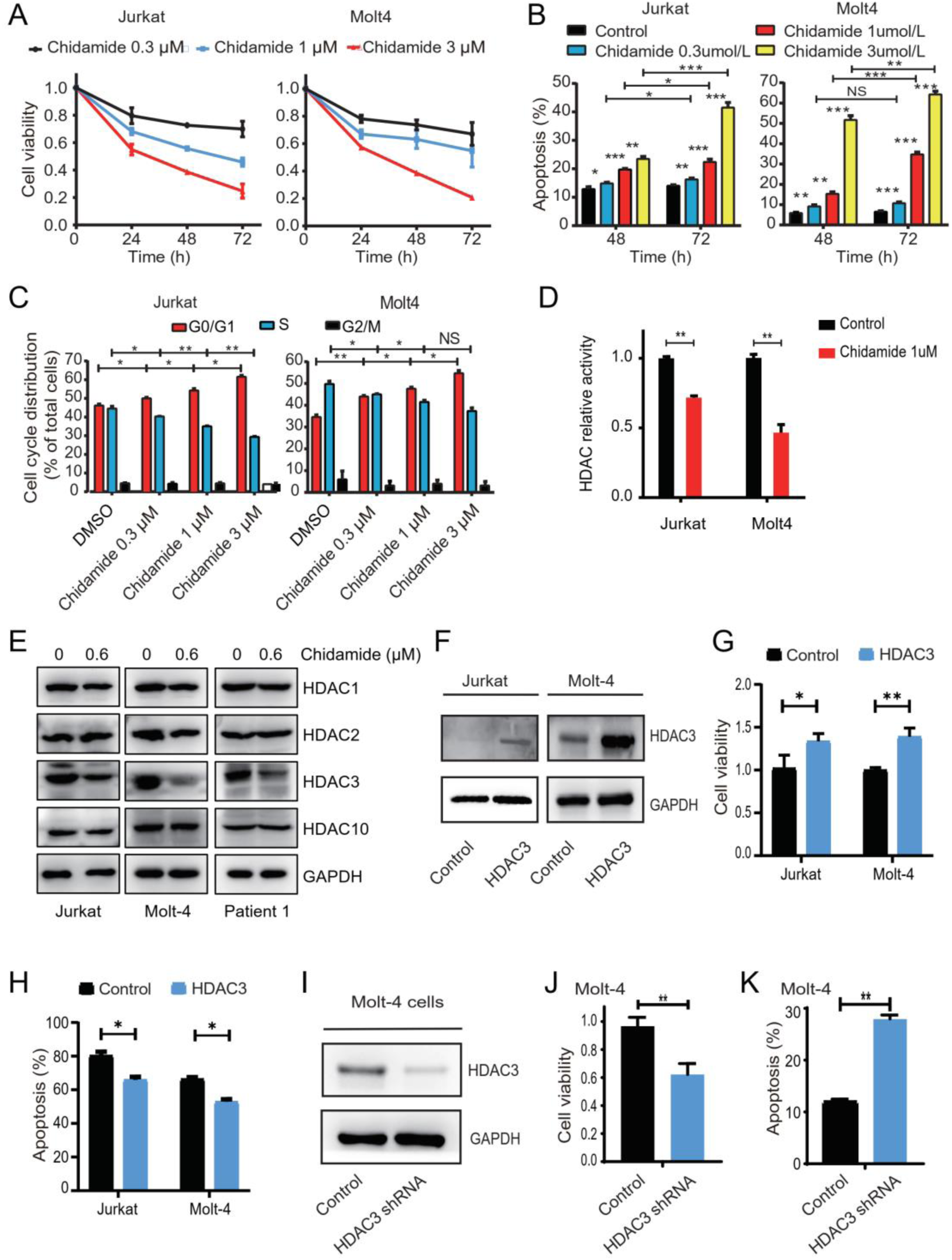
Chidamide significantly inhibited the growth of T-ALL cells via HDAC3. Jurkat and MOLT-4 cells were cultured with different concentrations of chidamide for 24, 48, and 72 h. **A** CCK-8 assays were used to assess cell viability. **B** Apoptosis was measured by flow cytometric analysis. Values are mean ± SD of the percentages of annexin V-positive cells in triplicate experiments. **C** Proportion of G0/G1 phase and S phase cells in response to incubation with chidamide for 24 h was measured with flow cytometry. **D** HDAC activity in chidamide-treated cells was significantly reduced compared to that in untreated cells in both cell lines. HDAC activity in control and chidamide-treated cells was measured by absorbance at 450 nm. **E** Western blot of HDAC3 in Jurkat, MOLT-4, and primary T-ALL cells treated with chidamide for 24 h. **F** Both Jurkat and MOLT-4 cells were transfected with overexpressing *HDAC3* plasmids or control plasmids and verified through Western blotting. The transfected cells were treated with 1 μM chidamide for 24 h. Overexpression of HDAC3 in response to chidamide is shown in **G** and **H**. **G** Cell proliferation was assessed through CCK-8 assays. **H** Evaluation of apoptosis by flow cytometry. Transfection with *HDAC3* shRNA was performed in Molt4 cells and verified through Western blotting (**I)**. The proliferation of the transfected cells was assessed **(J, K)**. **J** CCK-8 assays were used to assess cell viability. **K** Apoptosis was measured by flow cytometric analysis. Data represent three independent experiments. Results are shown as mean ± SD (*P < 0.05, **P < 0.01, ***P < 0.001, NS: P > 0.05).

Given that the pleiotropic effects of chidamide and its relevant molecular mechanism remained to be clarified, the global HDAC activity in both cell lines was evaluated. Upon treatment with chidamide, the total cellular deacetylase activity was significantly inhibited (Fig. 2D). This was consistent with a previous study on T-ALL[27]. The antitumor effect of HDACIs was initially considered to be dependent on their effect on histone acetylation. However, increasing evidence indicated that the biological effects of HDACIs are various[32]. Changes in expression levels of HDACs may also be necessary for the biological effects of HDACIs[32–34]. Therefore, we also checked the expression level of HDAC1, HDAC2, HDAC3, and HDAC10 upon treatment with chidamide (Fig. 2E and Figure S2A). The expression of HDAC3, but not HDAC1, HDAC2, or HDAC10, was dramatically downregulated after treatment with chidamide in both primary T-ALL specimens and cell lines. These findings suggested that the inhibitory effect of chidamide on T-ALL cells may be dependent on HDAC3.

### The inhibitory effect of chidamide on T-ALL cells was dependent on HDAC3

To determine whether chidamide-induced cell arrest of T-ALL cells was dependent on HDAC3, we first evaluated the effect of HDAC3 silencing on the viability of T-ALL cells. Next, we explored whether HDAC3 overexpression rescued the inhibitory effect of chidamide on T-ALL cells. First, *HDAC3* was overexpressed in both cell lines (Fig. 2F). Overexpression of *HDAC3* was found to rescue the decreased cell viability (Fig. 2G), decrease the percentage of apoptotic cells (Fig. 2H, and Figure S2B, C), and cause cell cycle arrest (Figure S2D,E,F) induced by chidamide. Then, the expression of *HDAC3* was knocked down in MOLT-4 cells (Fig. 2I). The cell proliferation was significantly suppressed in *HDAC3* knockdown cells (Fig. 2J). The percentage of apoptotic cells in the *HDAC3* knockdown cells was higher than that in control cells (Fig. 2K and Figure S2G). Moreover, *HDAC3* silencing was found to reduce the proportion of cells in the S phase and induced cell cycle arrest in the G0/G1 phase (Figure S2 H,I,J). Altogether, these data suggested that the inhibitory effect of chidamide on T-ALL cells was at least partially dependent on HDAC3.

### The TYK2-STAT1-BCL2 signaling pathway was inhibited by chidamide

To further explore the underlying mechanism of the inhibitory effect of chidamide on T-ALL cells, we performed RNA sequencing (RNA-seq) on chidamide-treated Jurkat and MOLT-4 cells. Cluster analysis of gene expression is shown in Fig. 3A and Figure S3A. Compared with untreated Jurkat cells, 2331 upregulated genes and 935 downregulated genes were identified in chidamide-treated Jurkat cells. There were 2478 upregulated genes and 806 downregulated genes in chidamide-treated MOLT-4 cells compared to untreated cells (Fig. 3A). In the above two cell lines, 983 co-differentially expressed genes were identified. And then, further Gene Ontology (GO) and Kyoto Encyclopedia of Genes and Genomes (KEGG) pathway enrichment analyses were performed (Fig. 3B and Figure S3B). Several signaling pathways, including the JAK-STAT signaling pathway, were significantly inhibited upon chidamide treatment (Fig. 3B). Within the JAK-STAT signaling pathway, the expression of *TYK2*, *STAT1*, and *BCL2* was significantly different (Fig. 3C). *HDAC3*, but not *HDAC1*, *HDAC2*, or *HDAC10*, was also found to be one of the most differentially expressed genes in both cell lines upon treatment with chidamide (Fig. 3C).

**Fig. 3.**
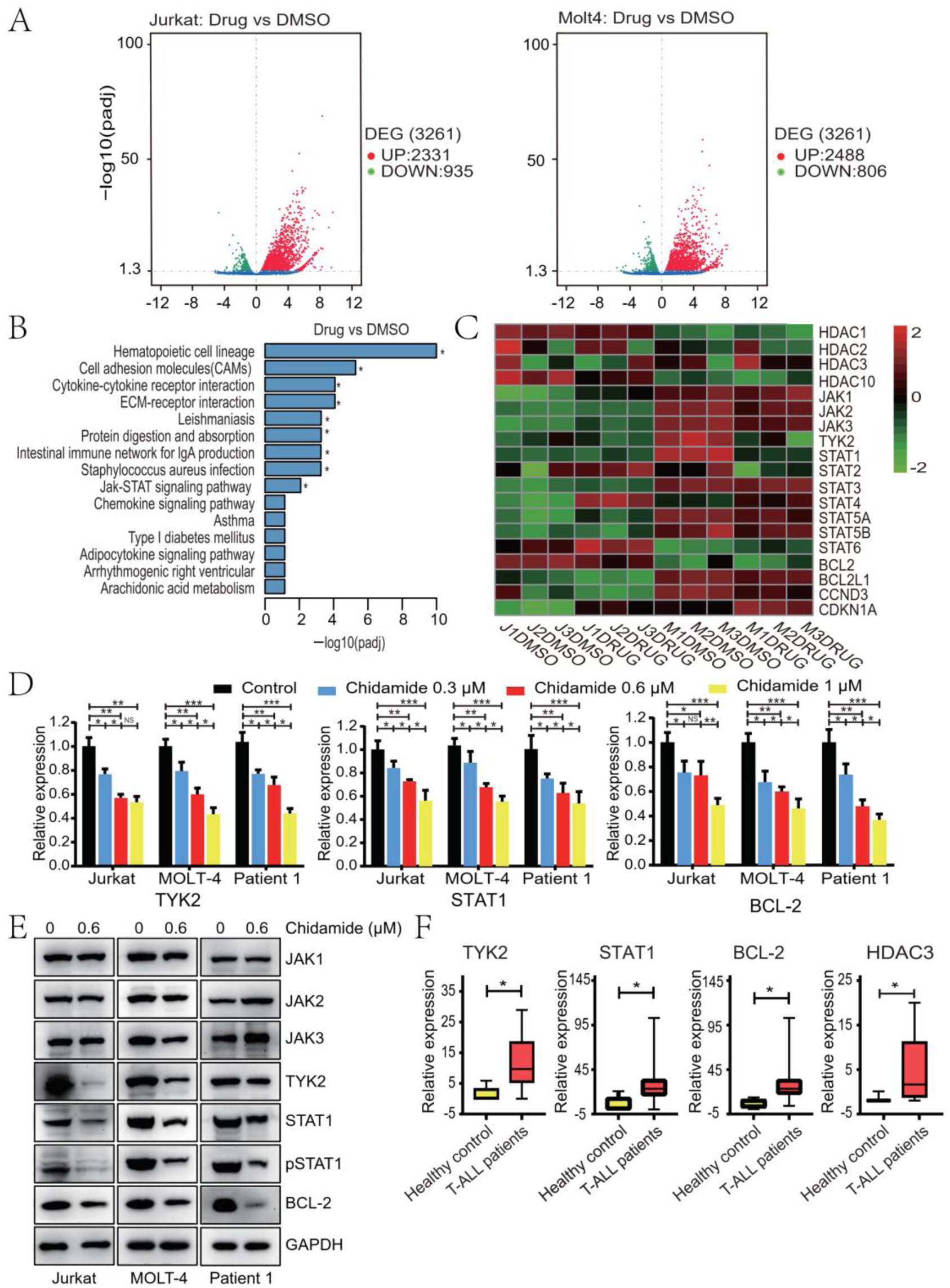
Differential gene and protein expression of T-ALL cells upon chidamide treatment. Total RNA isolated from T-ALL cell lines treated with DMSO or chidamide was subjected to RNA sequencing. **A** Volcano plot of differentially expressed genes defined using analysis of variance. **B** KEGG pathway analysis of differentially expressed genes. **C** Heat map of genes upon treatment with chidamide by analyzing the transcriptome. Differential gene expression was measured by RT-PCR **(D)** and Western blot **(E)** in both T-ALL cell lines and primary samples. **(F)** The mRNA expression levels of *TYK2*, *STAT1*, *BCL2*, and *HDAC3* of mononuclear samples from 29 patients with primary T-ALL and five heathy men were measured by RT-PCR. Data represent three independent experiments, and the results are shown as mean ± SD (*P < 0.05, **P < 0.01, ***P < 0.001).

To verify these results, we reviewed the differential expression of genes within the JAK-STAT signaling pathway in both cell lines and primary T-ALL samples. The mRNA and protein levels of TYK2, but not of JAK1–3, were both significantly inhibited upon chidamide treatment (Fig. 3D, E). The expression of STAT1, phosphorylated STAT1, and BCL2 was also clearly inhibited (Fig. 3D, E and Figure S3C). These data were in accordance with the RNA-seq results. HDAC has been reported to affect the growth of tumor cells by regulating the transcription of many oncogenes or tumor suppressor genes[35]. More importantly, the alteration of HDAC expression can protect tumor cells from genotoxic drugs[36]. The expression of *HDAC1*, *HDAC2*, *HDAC3*, and *HDAC10* genes upon chidamide treatment was examined (Fig. 2E). The protein level of HDAC3 was found to be significantly inhibited by chidamide. Given that the TYK2-STAT1-BCL2 signaling pathway has been reported to be constitutively activated in T-ALL cells[37], we also examined the expression level of the TYK2-STAT1-BCL2 signaling pathway in primary T-ALL cells. Compared with healthy controls, the mRNA level of *TYK2* and *STAT1* was also found to be upregulated (Fig. 3F). The expression level of *HDAC3* was found to be higher in primary T-ALL cells (Fig. 3F). All these results suggested that the TYK2-STAT1-BCL2 signaling pathway and HDAC3 were activated in T-ALL cells and that they could be inhibited by chidamide.

### Chidamide potently inhibits the growth of T-ALL cells via the TYK2-STAT1-BCL2 signaling pathway

First, we examined whether the growth of T-ALL cells was dependent on the TYK2-STAT1-BCL2 signaling pathway. *TYK2* was knocked down in Jurkat and MOLT-4 cells. The cell proliferation of both cell lines was significantly suppressed in *TYK2* knockdown cells (Fig. 4A). The percentage of apoptotic cells in *TYK2* knockdown cells was higher than that in the control cells (Fig. 4B, C). Moreover, *TYK2* silencing was found to reduce the proportion of cells in the S phase and induced cell cycle arrest in the G0/G1 phase (Fig. 4D, E and Figure S4A). We also examined the influence of *TYK2* knockdown on its downstream signaling pathway. The expression of STAT1, phosphorylated STAT1, and BCL2 was significantly inhibited in *TYK2* knockdown cells (Fig. 4F–I and Figure S4B–E). Taken together, our results consistently demonstrated that the survival of T-ALL cells was dependent on the TYK2-STAT1-BCL2 signaling pathway[37].

**Fig. 4.**
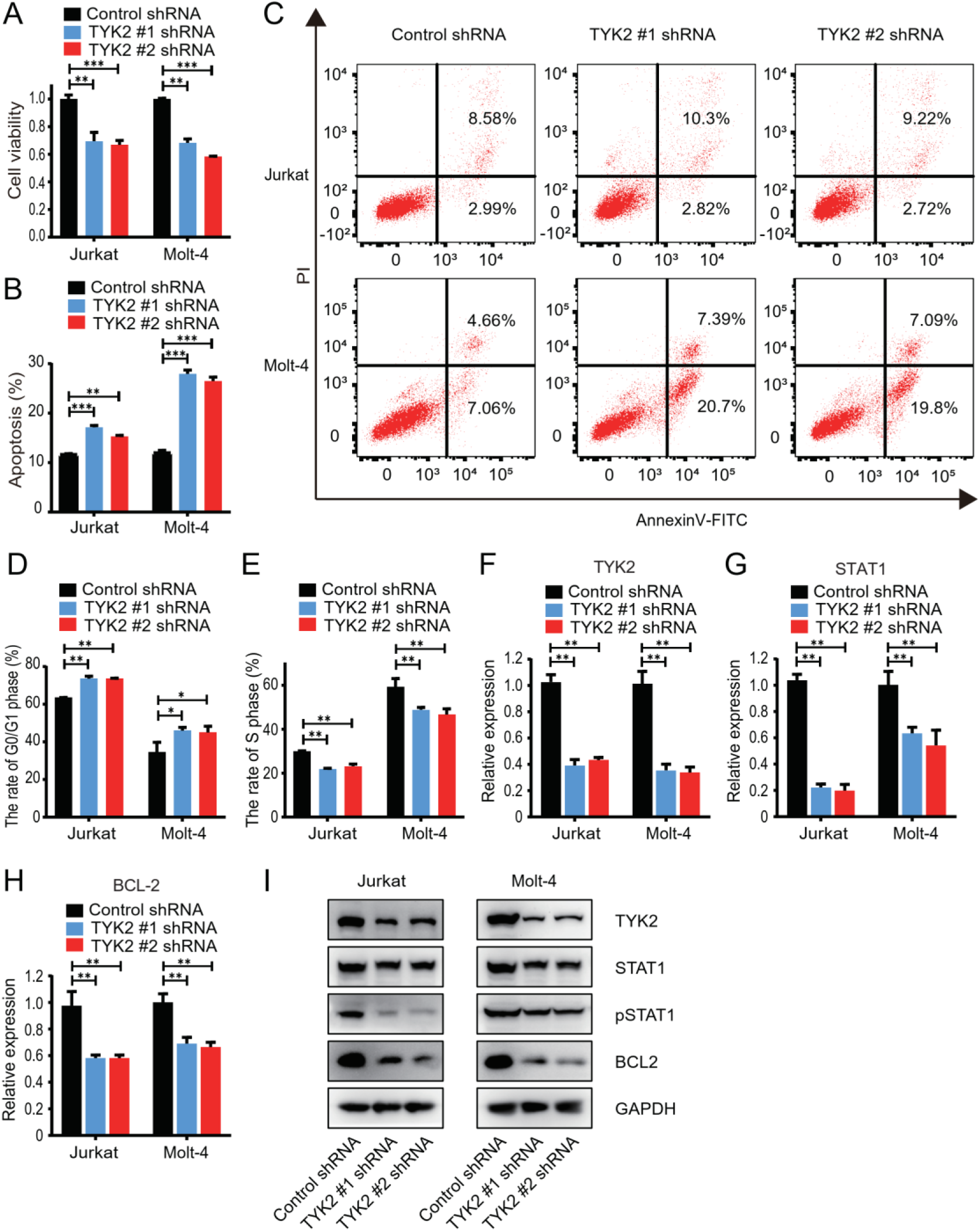
TYK2 silencing reduced the growth of T-ALL cells and suppressed the TYK2-STAT1-BCL2 signaling pathway. Transfection with TYK2 shRNA was performed in Jurkat and Molt4 cells. **A** Cell viability after transfection was assessed by CCK-8 assays. **B** Apoptosis results of flow cytometric analysis are mean ± SD percentages from triplicate experiments. **C** Representative results of apoptosis by flow cytometry. **D, E** The proportion of G0/G1 phase and S phase cells after TYK2 knockdown was measured with flow cytometry. **F–I** The expression of TYK2, STAT1, and BCL2 after transfection with TYK2 shRNA was measured by RT-PCR and Western blot. Data represent three independent experiments, and the results are shown as mean ± SD (*P < 0.05, **P < 0.01, ***P < 0.001).

Next, we explored whether chidamide-induced cell arrest of T-ALL cells was dependent on TYK2. As expected, overexpression of *TYK2* was found to rescue the decreased cell viability (Fig. 5A), decrease the percentage of apoptotic cells (Fig. 5B, C and Figure S5A), and alleviate cell cycle arrest (Fig. 5D–F and Figure S5B) induced by chidamide. The expression of STAT1, phosphorylated-STAT1, and BCL2 was found to be upregulated in cells with *TYK2* overexpression (Fig. 5G–J and Figure S5C–E). Altogether, our data supported that the inhibitory effect of chidamide on T-ALL cells was at least partially in a TYK2-dependent manner.

**Fig. 5.**
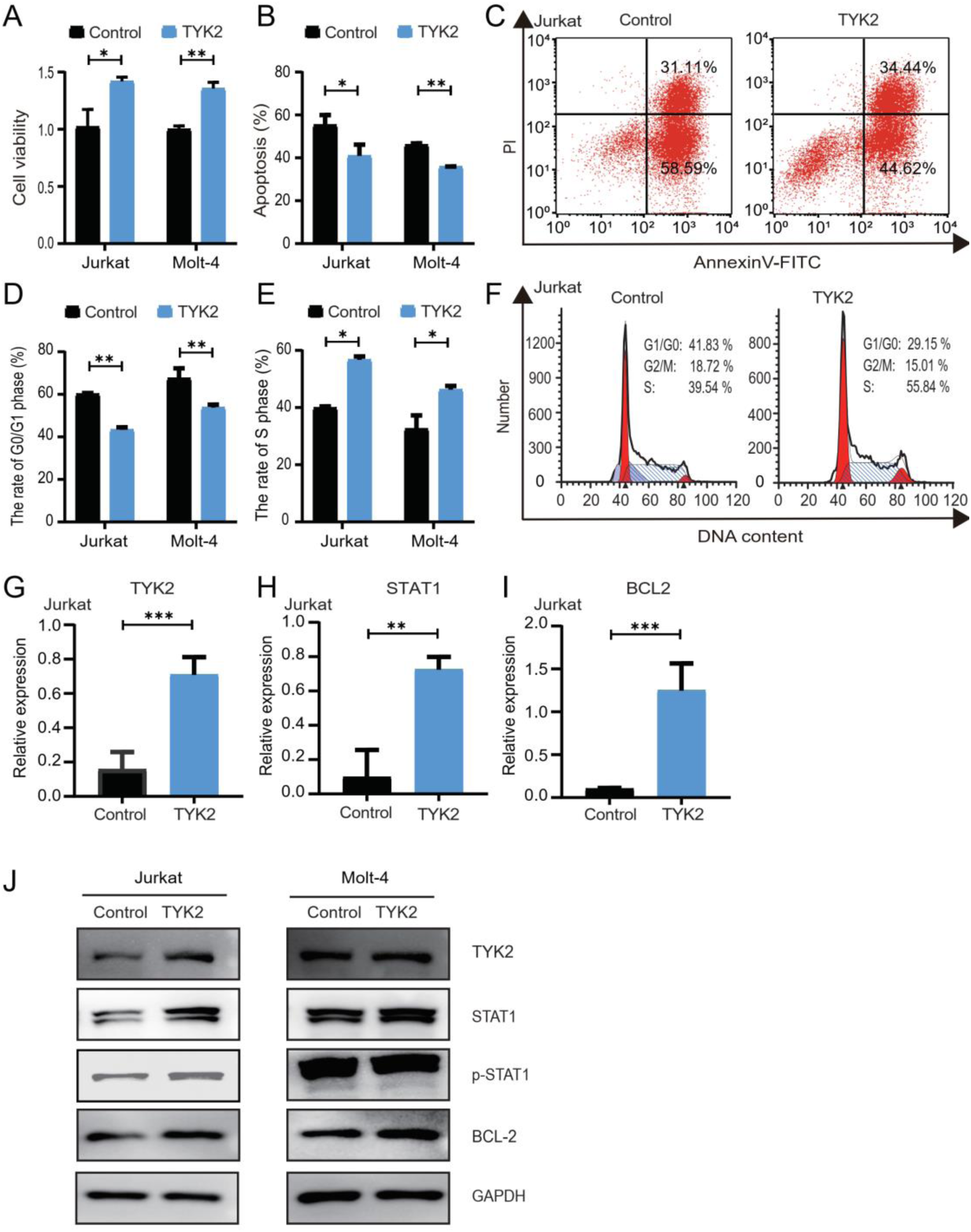
Overexpression of TKY2 rescued the inhibitory effect of chidamide via the TYK2-STAT1-BCL2 signaling pathway. Jurkat and MOLT-4 cells were transfected with plasmids overexpressing TYK2 or control plasmids. Transfected cells were treated with 1 μM chidamide for 24 h. **A** Cell viability was assessed by CCK-8 assays. **B** Apoptosis results of flow cytometric analysis are mean ± SD percentages of triplicate experiments. **C** Representative results of apoptosis by flow cytometry. **D–F** The proportion of G0/G1 phase and S phase cells after TYK2 overexpression. **G–J** The expression of TYK2, STAT1, and BCL2 after transfection was measured by RT-PCR and Western blot (*P < 0.05, **P < 0.01, ***P < 0.001).

### HDAC3 bound to TYK2 and activated the TYK2-STAT1-BCL2 signaling pathway

Given that chidamide inhibited the growth of T-ALL cells and that overexpression of *HDAC3* or *TYK2* at least partially rescued the inhibitory effect of chidamide, we sought to explore the impact of HDAC3 expression on the TYK2-STAT1-BCL2 signaling pathway and the interaction between HDAC3 and TYK2. We found that *HDAC3* silencing decreased the expression of TYK2, STAT1, phosphorylated STAT1, and BCL2 (Fig. 6A–D). Similarly, overexpression of *HDAC3* led to the upregulation and activation of the TYK2-STAT1-BCL2 signaling pathway (Fig. 6E–H and Figure S6A–D). Then, we performed co-immunoprecipitation to verify the protein interaction between HDAC3 and TYK2. As expected, HDAC3 directly bound to TYK2 (Fig. 6I and Figure S6E). To further explore the interaction between HDAC3 and TYK2 and whether HDAC3 was responsible for TYK2 phosphorylation, co-immunoprecipitation was performed to verify the HDAC3 and TYK2 interaction in HEK293T cells co-expressing HDAC3 and TYK2. The presence of a high level of HDAC3 was associated with a higher level of phosphorylated TYK2 and thus lower acetylated TYK2 compared to that in cells expressing TYK2 alone, which might result from endogenous HDAC3 (Fig. 6J). Taken together, these results suggested that HDAC3 binds to TYK2 and contributes to the activation of the TYK2-STAT1-BCL2 signaling pathway.

**Fig. 6.**
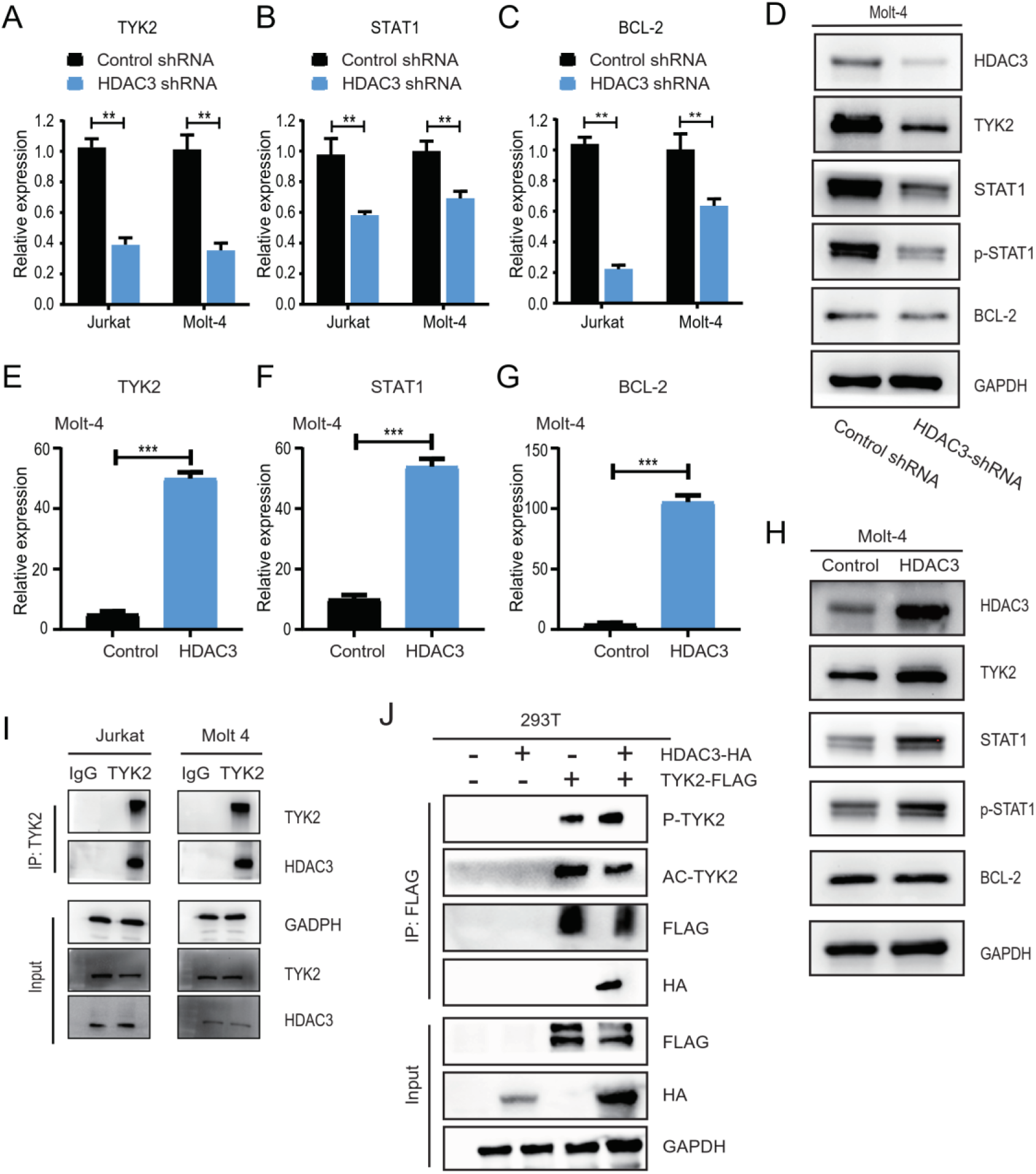
The inhibitory effect of chidamide on T-ALL cells was dependent on the HDAC3-TYK2-STAT1-BCL2 signaling pathway. **A–D** Jurkat and MOLT-4 cells were transfected with HDAC3 shRNA. The expression of TYK2 **(A)**, STAT1 **(B)**, and BCL2 **(C)** after transfection was measured by RT-PCR. **D** The expression of TYK2, STAT1, p-STAT1, and BCL2 after transfection with HDAC3 shRNA was measured by Western blot. **E-H** Molt-4 cells were transfected with overexpressing HDAC3 plasmids or control plasmids. The mRNA expression of TYK2 **(E)**, STAT1 **(F)**, and BCL2 **(G)** after transfection was measured by RT-PCR. **H** The expression of TYK2, STAT1, p-STAT1, and BCL2 after transfection was evaluated by Western blot. **I** Co-immunoprecipitation of TYK2 and HDAC3 was performed using Jurkat and Molt-4 cells. **J** Co-immunoprecipitation of TYK2 and HDAC3 was performed in 293 T cells. Data represent three independent experiments, and the results are shown as mean ± SD (**P < 0.01, ***P < 0.001).

### Chidamide and a TYK2 inhibitor synergistically inhibited the growth of T-ALL cells both in vitro and in vivo

To further validate whether there was a synergistic inhibitory effect between chidamide and TYK2 inhibition in T-ALL cells, deucravacitinib, a highly selective inhibitor of TYK2, was used. The proliferation of T-ALL cells was profoundly inhibited by the combination treatment of chidamide and deucravacitinib (Fig. 7A). Compared with a single reagent, the percentage of apoptotic cells was significantly higher in the cells treated with the combination treatment of chidamide and deucravacitinib (Fig. 7B and Figure S7A). Then, a xenograft mouse model was established in NOD/SCID mice by subcutaneously inoculating the mice with wild-type Jurkat cells (total = 24) (Fig. 7C–G). The mice were randomized into four groups (n = 6). They were treated with chidamide, deucravacitinib, chidamide with deucravacitinib, or a PBS control. The volume and weight of the tumors in the combination group were less than those of tumors in the other groups (Fig. 7C–F). Moreover, the expression of phosphorylated STAT1, BCL2, and Ki-67 in the combination group was lower than that in the other groups (Fig. 7G and Figure S7B,C). These results indicated that chidamide and a TYK2 inhibitor synergistically inhibited the growth of T-ALL cells.

**Fig. 7.**
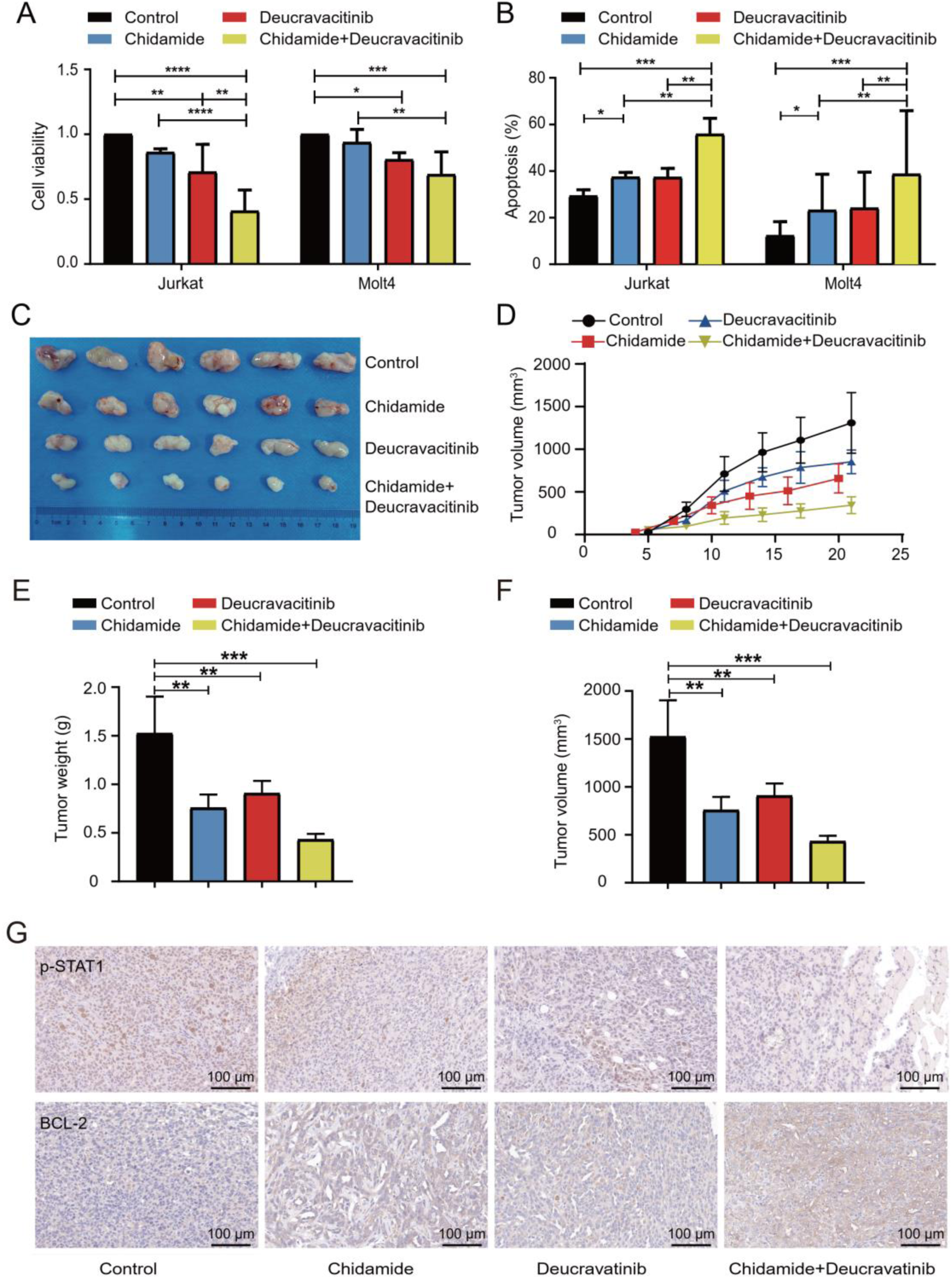
Chidamide and a TYK2 inhibitor synergistically inhibited the growth of T-ALL cells both in vitro and in vivo. Jurkat and Molt-4 cells were incubated with 1 μM chidamide or 4 μM deucravacitinib as a monotherapy or with the combination of chidamide and deucravacitinib for 36 h. **A** CCK-8 assays were used to assess cell proliferation. **B** Flow cytometry was used to analyze cell apoptosis. Jurkat cells (1 × 10^7^ cells) were implanted into NOD/SCID mice. The mice were randomly divided into four groups (six mice in each group). The mice in the chidamide-treated group were intragastrically administered with chidamide three times a week for 2 weeks. The mice in the deucravacitinib-treated group were intragastrically administered deucravacitinib twice daily for 2 weeks. The mice in the control group were treated with both PBS and normal saline as a control. **C** The volume of each tumor was measured every 3 days. The tumor volume was calculated using the formula: V = 0.5 × length × width2. **D** A visual analysis of tumors harvested from mice. **E, F** The measurement of xenograft tumor volume and weight. **G** Representative expression of p-STAT1, BCL2 in tumor sections by immunohistochemistry. Data represent three independent experiments and are expressed as mean values ± SD (*P < 0.05, **P < 0.01, ***P < 0.001, NS: P > 0.05).

## Discussion

In this study, our results showed that chidamide-based combination regimens significantly improved the response rates of 28 patients with R/R T-ALL. The overall response and CR rate were 68% and 57% with the combined regimen, respectively. Because of this, quite a few patients in our study were able to receive allo-HSCT and achieved longer survival. Moreover, our study demonstrated that chidamide potently inhibited the growth of T-ALL cells via the HDAC3-TYK2-STAT1-BCL2 signaling pathway. We found that the HDAC3-TYK2-STAT1-BCL2 signaling pathway was activated in T-ALL cells. Our results further demonstrated that HDAC3 bound to TYK2 and contributed to the activation of the TYK2-STAT1-BCL2 signaling pathway in T-ALL cells. Our finding is the first to implicate HDAC3 in the TYK2-STAT1-BCL2 signaling pathway. Considering that effective inhibitors against TYK2 and BCL2 are far from optimal for clinical application, our study suggested that HDAC3 can work as a potential alternative therapeutic target to inhibit the TYK2-STAT1-BCL2 signaling pathway in T-ALL.

The clinical outcome for adult patients with R/R T-ALL is still very poor, and it remains a major challenge for most hematologists. Currently, there is no widely accepted salvage regimen for this group of patients. The efficacy of combination chemotherapy is far from satisfactory. Despite the limited efficacy, nelarabine has been approved for patients with R/R T-ALL[38]. Allo-HSCT remains the only potential curative option for longer survival[39]. However, uncontrolled disease status is a known risk factor for the failure of allo-HSCT. An effective salvage regimen to achieve CR, followed by allo-HSCT, is considered to be the best strategy for patients with R/R T-ALL. Hence, new salvage strategies are urgently needed for this group of patients. Both the genetic features and epigenetic alterations in T-ALL have been thoroughly studied[11, 13, 14]. Unfortunately, the results of preclinical and clinical studies assessing inhibitors or drugs against oncogenic drivers within the Notch or mTOR signaling pathways are still somewhat disappointing[40, 41]. Given that there are also deregulated HDAC alterations in T-ALL[14], we first tested the efficacy of chidamide combined with chemotherapy. The CR rates reached as high as 57% using the combined regimen. Currently, the CR rate of conventional salvage chemotherapy is less than 50%[30]. These encouraging clinical outcomes prompted us to investigate the underlying mechanism of the inhibitory effect of chidamide on T-ALL cells.

We tested the inhibitory effects of chidamide on T-ALL cells. In line with previous studies[33], we found that chidamide significantly inhibited the proliferation of T-ALL cells and induced apoptosis and cell cycle arrest in vitro. Given the fundamental role of histone acetylation in chromatin remodeling and transcription, the antitumor effect of HDACIs was initially considered to be through regulation of gene expression via direct histone hyperacetylation. However, increasing evidence indicates that many non-histone proteins are also regulated by HDACs. The biological effects of HDACIs are various and some of them are associated with non-histone proteins[32]. Altered gene expression could be the direct effect of HDACIs through histone hyperacetylation in certain circumstances. Changes in activity or expression levels of HDACs may also be necessary for the biological effects of HDACIs[32–34]. Next, we checked the expression level of HDACs upon treatment with chidamide in T-ALL cells. Chidamide specifically downregulated the expression of HDAC3, but not of HDAC1, HDAC2, or HDAC10, in both primary T-ALL specimens and cell lines. Consistent with a previous study[14], the expression level of HDAC3 in primary T-ALL cells was also found to be higher than that in normal bone marrow samples. Previous studies proposed that overexpression of HDACs was vital for the maintenance of genome stability and DNA damage control in tumor cells, thus protecting cells from exogenous genotoxic insults[36, 42]. Similar to the role of HDAC3 reported in multiple myeloma and cutaneous T-cell lymphoma[43, 44], our study demonstrated that knockdown or pharmacological inhibition of *HDAC3* inhibited the growth of T-ALL cells and induced apoptosis and cell cycle arrest. Overexpression of *HDAC3* rescued the inhibitory effect of chidamide. Taken together, our study suggests that chidamide inhibited the growth of T-ALL cells by downregulating the expression level of HDAC3, highlighting HDAC3 as an attractive and potential target in T-ALL.

Given the increasing number of HDAC substrates, the biological effects of HDACIs are found to be broader and more complicated than originally thought. Accumulating evidence suggests that it is unlikely that only a certain signaling pathway accounts for the effects of all HDACIs in all cell types[34]. Thus, it is logical and reasonable to postulate that the effects of HDACIs can be cell type-specific. Additionally, there is increasing data suggesting that the biological effects of structurally different HDACIs can be different even in the same cell types[24]. To obtain a better understanding of the molecular pathways that are engaged to mediate the anticancer effects of chidamide in T-ALL, we performed RNA-seq in chidamide-treated and DMSO-treated T-ALL cells. The TYK2-STAT1-BCL2 signaling pathway was found to be significantly suppressed upon treatment with chidamide. Considering that a previous study demonstrated that the survival of T-ALL cells was dependent on the TYK2-STAT1-BCL2 signaling pathway[37], our study confirmed that chidamide inhibited the growth of T-ALL cells via the TYK2-STAT1-BCL2 signaling pathway. We further showed that HDAC3 bound to TYK2 and contributed to the activation of the TYK2-STAT1-BCL2 signaling pathway. To the best of our knowledge, our study is the first that demonstrates the connection of HDAC3 with the TYK2-STAT1-BCL2 signaling pathway. Taken together, our results suggest that the inhibitory effects of chidamide on T-ALL cells are dependent on the HDAC3-TYK2-STAT1-BCL2 signaling pathway.

Our study demonstrated that knockdown of *TYK2* inhibited the proliferation of T-ALL cells and reduced the expression level of STAT1, phosphorylated STAT1, and BCL2. This is in line with a previous study that demonstrated that the survival of T-ALL cells was dependent on the TYK2-STAT1-BCL2 signaling pathway[37]. Importantly, our study further demonstrated that overexpression of *TYK2* rescued chidamide-induced growth arrest of T-ALL cells. Furthermore, knockdown of *HDAC3* was also found to downregulate the expression of TYK2, STAT1, phosphorylated STAT1, and BCL2, thus inducing growth arrest of T-ALL cells. Accordingly, overexpression of *HDAC3* activated and upregulated the expression of the TYK2-STAT1-BCL2 signaling pathway, presumably rescuing the inhibitory effect of chidamide on T-ALL cells. All these data collectively further demonstrate the important role of the TYK2-STAT1-BCL2 signaling pathway for the survival of T-ALL cells. Although great effort is ongoing to develop more potent TYK2 and BCL2 inhibitors and evaluate their clinical role in T-ALL[45–47], none are available for clinical use. Therefore, researchers are developing alternative strategies to inhibit the TYK2-STAT1-BCL2 signaling pathway. Akahane et al. proposed to promote or accelerate the degradation of TYK2 kinase by HSP90 inhibition and induce apoptotic cell death in T-ALL[48]. In this study, our results demonstrated that HDAC3 bound directly to TYK2, contributing to the upregulation and activation of the TYK2-STAT1-BCL2 signaling pathway. Furthermore, knockdown or pharmacological inhibition of HDAC3 with chidamide suppressed the TYK2-STAT1-BCL2 signaling pathway and induced growth arrest of T-ALL cells. Collectively, these findings suggest that HDAC3 has the potential to be an alternative therapeutic target to inhibit the TYK2-STAT1-BCL2 signaling pathway in T-ALL.

In summary, our study suggests that the combination of chidamide and chemotherapy is a promising treatment for patients with relapsed/refractory T-ALL. Nevertheless, this combined regimen still needs to be further confirmed in large prospective clinical trials. Mechanistically, we found that chidamide significantly inhibited the growth of T-ALL cells via the HDAC3-TYK2-STAT1-BCL2 signaling pathway. HDAC3 directly bound to TYK2 and contributed to the activation of the TYK2-STAT1-BCL2 signaling pathway in T-ALL cells. This finding demonstrated the association of HDAC3 with the TYK2-STAT1-BCL2 signaling pathway, suggesting that HDAC3 may become an alternative and potential therapeutic target to inhibit the TYK2-STAT1-BCL2 signaling pathway in T-ALL.

## Data Availability

All data generated or analyzed during this study are included in this published article. The datasets used and/or analyzed during the current study are available from the corresponding author upon reasonable request.

## Abbreviations

HDAC3: histone deacetylase 3
T-ALL: T-cell acute lymphoblastic leukemia/lymphoma
TYK2: tyrosine kinase 2
p-TYK2: phospho-TYK2
STAT1: signal transducer and activator of transcription 1
p-STAT1: phospho-STAT1
BCL2: B-cell lymphoma-2
B-ALL: B-cell acute lymphoblastic leukemia
HDAC: histone deacetylase
HDAC1: histone deacetylase 1
HDAC2: histone deacetylase 2
HDAC10: histone deacetylase 10
ALL: acute lymphoblastic leukemia/lymphoma
CAR-T: chimeric antigen receptor T-cell immunotherapy
NOTCH: neurogenic locus notch homolog protein
WNT: wingless
MYC: myc proto-oncogene
R/R: relapsed/refractory
T-LBL: T-cell lymphoblastic lymphoma
ETP-ALL: early T-cell precursor acute lymphoblastic leukemia
CR: complete remission
PR: partial response
PD: progressive disease
NR: no response
ORR: overall response rate
DMSO: dimethyl sulfoxide
ATCC: America type culture collection
PBS: phosphate buffered saline
CCK-8: Cell Counting Kit-8
OD: optical density
PI: propidium iodide
RNase A: ribonuclease A
FITC: fluorescein isothiocyanate
RPKM: reads per kilobase per million mapped reads
PCR: polymerase chain reaction
RT-PCR: real-time PCR
cDNA: complementary DNA
shRNA: short hairpin RNA
RIPA: radio immunoprecipitation assay
SDS-PAGE: SDS-polyacrylamide gel electrophoresis
PVDF: polyvinylidene difluoride
NOD/SCID: non obese diabetes/server combined immune-deficiency
IACUC: Institutional Animal Care and Use Committee
SD: standard deviation
allo-HSCT: allogeneic hematopoietic stem cell transplantation
HDACIs: histone deacetylase inhibitors
RNA-seq: RNA sequencing
GO: Gene Ontology
KEGG: Kyoto Encyclopedia of Genes and Genomes
JAK: janus kinase
STAT: signal transducer and activator of transcription
mRNA: messenger RNA
mTOR: mammalian target of rapamycin
HSP90: heat shock protein 90
IRBs: institutional review boards
WBC: white blood cell
Hb: Hemoglobin
BM: bone marrow
CNS: central nervous system
NA: not applicable
OS: overall survival
PFS: progression-free survival
Co-IP: co-inmunoprecipitation

## Declarations

## Ethics approval and consent to participate

Samples were collected after obtaining written informed consent according to the protocols approved by the institutional review boards (IRBs) of Chinese PLA General Hospital. All experiments were conducted in accordance with the Declaration of Helsinki.

## Consent for publication

The authors declare that they consent for publication.

## Authors’ contributions

LPD, DHL, CJG, and YX designed the study and initiated this work. Data were obtained by ZYG, YFJ, YCL, HW, LLW, XWZ and QYL. Experiments were performed by ZYG, YFJ, YCL, HW, LLW, XWZ and QYL. All statistical analyses were performed by ZYG, YFJ, YCL, HW, LLW. ZYG wrote the paper. All the authors were involved in the interpretation of the results; read, provided comments, and approved the final version of the manuscript; had full access to the data in the study; and take responsibility for the accuracy of the data analysis.

## Acknowledgements

We thank all the faculty members who participated in this study.

